# A conserved grain-associated immunosuppressive niche in Sudanese patients with mycetoma

**DOI:** 10.64898/2026.04.09.26350374

**Authors:** Mohamed Osman, Helen Ashwin, Grant Calder, Peter O’Toole, Sahar M. Bakhiet, Ahmed M. Musa, Paul M. Kaye, Ahmed Hassan Fahal

## Abstract

Mycetoma is a neglected tropical disease caused by various bacterial and fungal pathogens that has a significant health impact across a broad geographically defined “mycetoma belt” spanning South America, Africa and Asia. Histologically, mycetoma is characterised by invasive and destructive granuloma development in the skin, deep tissues and bone, leading to tissue destruction, deformities and high morbidity. The presence of macroscopic, highly compacted pathogen microcolonies, or “grains,” is a key diagnostic feature, and the formation of grains supports pathogen persistence and disease chronicity. However, there is a paucity of information on immune responses in mycetoma patients and on the relative importance of phylogeny and/or grains in establishing the local immune landscape. Here, we used spatial proteomics to examine the distribution of 43 immune-related proteins in surgical biopsies from 11 patients with mycetoma of bacterial (actinomycetoma; *Actinomadura pelletierii* and *Streptomyces somaliensis;* n=6) and fungal (eumycetoma; *Madurella mycetomatis;* n=5) origin. Using mixed-effects modelling, an exploratory analysis across species and pathogen classes revealed few significant differences in immune marker expression. In contrast, and independently of pathogen class, the cellular infiltrate closest to grain boundaries had higher per-cell expression of CD66b^+^ neutrophils, ARG1, and VISTA. The preferential accumulation of CD66b^+^ARG1^+^VISTA^+^ cells at grain boundaries was confirmed by quantitative immunofluorescence analysis. Hence, the local tissue microenvironment surrounding the mycetoma grain represents a specialised immunosuppressive niche, with parallels to the tumour microenvironment.

The study was prospectively registered on ClinicalTrials.gov (NCT04401969).

**Author summary:** Mycetoma is a devastating disease found in tropical countries that causes high levels of tissue damage, often leading to amputation of affected limbs. Various bacterial and fungal pathogens cause the disease, but this does not affect how the disease appears. A common feature of mycetoma that helps doctors distinguish it from other diseases is the presence of “grains” in the tissues, which are discharged through the open sinuses. Under the microscope, they appear as colonies of the pathogen, together with host cell debris. It has been speculated that these grains allow the pathogen to resist immune attack and therefore contribute to the chronic nature of the infection. To understand how this may occur, we used spatial profiling and immunohistology to map the locations of multiple immune markers in the tissues of patients diagnosed with mycetoma. Our results show that phagocytic cells (neutrophils) in closest proximity to grains express molecules associated with immunosuppression. Similar neutrophils are found in close association with tumour cells in patients with cancer, where they inhibit host immunity and reduce the effectiveness of anti-tumour drugs. We speculate that these cells may play a similar role in mycetoma patients.

## Introduction

Mycetoma, recently recognised by the WHO as a neglected tropical disease, is a chronic, potentially disfiguring subcutaneous granulomatous inflammatory disease [1–3]. Mycetoma is reported worldwide, but notably in a geographically defined “mycetoma belt,” and affects the poorest of the poor at the individual, family, community, and health system levels [4–6]. Mycetoma is caused by >70 species of true fungi (eumycetoma) or actinomycete bacteria (actinomycetoma), but the disease presentation is broadly similar across pathogen classes [2, 7, 8].

Diagnostically and histologically, mycetoma is associated with the presence of “grains”, compacted microcolonies of organisms associated with host cell debris and extracellular matrix components [2, 9–12]. Grain morphology and ultrastuctural characteristics can be used to classify disease by pathogen class [2, 7, 10, 13, 14], and the triad of a painless subcutaneous mass, multiple sinuses and the sero-purulent discharge that contains grains is a mycetoma characteristic [2]. The histological response has been characterised in patients with both eumycetoma and actinomycetoma and supports a histopathological picture that is associated with both myeloid (predominantly neutrophil) and lymphoid infiltration [15–19]. In eumycetoma, grading based on the presence of neutrophils, granuloma-associated giant cells, and necrosis has been proposed [16]. However, studies on specific immune cells populations and their products remain cursory, with a focus on T cell (TH1 and TH2) -associated cytokines, IL-1 family cytokines and mediators of neutrophil recruitment and activation, such as IL-17 and MMP9 [15, 16, 19–24]. Such tissue responses are likely shaped by responses to the pathogen and to tissue destruction, as well as by host genetics [24, 25].

Presently, treatment options for eumycetoma patients are limited and characterised by low cure and high recurrence rates [26–28], despite most causative fungi being susceptible to many antifungal drugs [29–32]. For actinomycetoma, a combination of antibacterials is available, but cure requires extensive treatment regimens that limit compliance [27]. Surgical intervention is required for the treatment of most eumycetoma patients, with unpredictable recurrence rates [33]. Although it is likely that chronicity and treatment response are intimately linked through the microenvironmental control of immune responses [34], this has not been formally explored in the context of mycetoma.

Here, we conducted an exploratory study to investigate the value of applying digital spatial profiling to study the mycetoma granuloma microenvironment. Using a 43-plex protein panel that features multiple immune-oncology targets, our analysis revealed only minor differences in global tissue response between eumycetoma and actinomycetoma patients. However, we found that CD66b^+^ARG1^+^VISTA^+^ cells were abundant at grain boundaries and shape an immunoregulatory landscape that may promote chronicity and limit drug effectiveness.

## Methods

### Ethics statement

The study was conducted in accordance with the principles of the Declaration of Helsinki. The Ethical Review Committee of Soba University Hospital, University of Khartoum and the Department of Biology Ethics Committee, University of York (Ref: MO201903) gave ethical approval for this work. All patients provided written consent for the use of diagnostic surgical biopsies in research aimed at understanding mycetoma pathogenesis. The study protocol did not impact the standard of care.

### Patients

This observational clinical study (ClinicalTrials.gov id: NCT04401969) was conducted at the Mycetoma Research Centre, Soba University Hospital, University of Khartoum, Khartoum, Sudan, with tissue analysis conducted at the University of York, UK. The study originally included 28 patients, and the diagnosis was confirmed by clinical examinations, culture of grains and histopathological examination of material obtained by a surgical biopsy. True-Cut surgical biopsies from all patients were taken as part of routine diagnostic procedures, fixed in 10% formalin for 24 hours, and processed to Formalin Fixed Paraffin Embedded (FFPE) sections. After staining with Haematoxylin and Eosin (H&E), 11 patients (six with actinomycetoma and five with eumycetoma) were selected for further study based on the ready identification of grains in FFPE sections. These patients were classified as having actinomycetoma caused by either *Actinomadura pelletieri* (n=4; P1, 4, 7, and 21) or S*treptomyces somaliensis* (n=2; P2 and 25), and eumycetoma caused by *Madurella mycetomatis* (P8, 17, 24, 27, and 28).

### GeoMx Digital Spatial Profiling

Spatial protein profiling on 5µm thick sections from FFPE tissue biopsies were performed using the NanoString GeoMx DSP™ assay system (Bruker). A profiling panel of 43 immune-oncology proteins together with housekeeping proteins and isotype controls were assessed. All staining, imaging, probes hybridisation and counting were done as described in the manufacturer protocols. Using the GeoMx DSP Control Center software (v2.4) two regions of interest (ROIs) with a 300µm diameter were manually created surrounding grains (called “Near”) and two more were created 200-300µm away from any grain perimeter (called “Far”) equalling four ROIs per patient and forty-four ROIs in total. UV light was used to release identifying bar codes from the ROI area which were sequentially collected for later identification and counting using the NanoString nCounter system. Digital count data were analysed using GeoMx Data analysis Suite (v2.4) where counts are normalised to positive External RNA Controls Consortium (ERCC) controls and to nuclei counts. ROIs with abnormal levels of hybridisation or housekeeping (HK) protein expression were removed from the analysis. Target proteins were only retained for analysis if counts were 3x the geometric mean of the isotype controls in >10% of ROIs. HK proteins were also excluded from analysis as their expression may vary under inflammatory conditions. Data for the 32 proteins passing QC and thresholding were exported for further analysis in R (see Statistical Analysis) and analysed as normalised counts using linear mixed modelling, adjusting for patient ID (repeat measures), pathogen class, and position (Near vs Far).

### Immunohistology

4µm sections from FFPE blocks were used for all analyses. Slides were heat fixed at 50 °C for 10 minutes in an oven. Paraffin was removed from the sections by sequential washes: 2x 5 minutes in Xylene, 3 minutes in 95% alcohol, 3 minutes in 70% alcohol, and then water for rehydration. Antigens were unmasked under high pressure in citrate buffer using a high-pressure cooker for 15 minutes. Following three washes with PBST (PBS, 0.05% Tween 20), Slides were blocked with the background suppressor TrueBlack® IF Background suppressor 23012A (Biotium) for 30 min at room temperature. Primary antibodies were diluted in blocking buffer(TrueBlack® IF Blocking buffer 23012B, Biotium), and incubated overnight at 4^°^C. Sections were then probed with the following antibodies; CD68 (ab955, 1:100) detected by Goat anti-Mouse AF546 (1:500), CD4 (ab133616, 1:100) detected by Goat anti-rabbit AF488 (1:400) or Donkey anti-rabbit Dy650 (1:500), CD8 (Biolegend 372902, 1:100) detected by Goat anti-Mouse AF546 (1:500), CD3 (Ab5690, 1:100) detected by Goat anti-rabbit AF488 (1:400) or Donkey anti-rabbit Dy650 (1:500), Arg1 (Invitrogen H1D5, 1:200) detected by Goat anti mouse F(ab)2 AF555 (Invitrogen A21425,1:500), CD66b AF647 (Biolegend 392912, 1:50) detected by Mouse IgG1 AF647 (Biolegend 400130, 1:500), VISTA (Cellsignal 64953S; 1:100) detected by Donkey anti-Rabbit CF750 (Biotium 20298, 1:500), CD20 Dy650 (Novus NBP-47840C, 1:100), CD56 Dy650 (Novus NBP-33132C, 1:100), and CD66b AF647 (Biolegend 392912, 1:20). Intermediate blocking using Mouse IgG (ab37355, 1:400) was included where necessary. Sections were counterstained with DAPI and mounted in FluoromountG mounting medium. Images were acquired using Zeiss AxioScan.Z1 slide scanner. Identical exposure times and threshold settings were used for each channel on all sections of similar experiments. Quantifications of cell types based on marker expression were performed using StrataQuest Analysis Software (TissueGnostics) within manually curated ROIs extending in 100um steps from grain perimeters (<100um, 101-200um, and 201-300um).

### Statistical Analysis

DSP protein expression data were analysed using linear models implemented in the *limma* framework, using log2-transformed normalised counts (log2[counts + 1]). To account for repeated measurements per patient, patient identity was modeled as a blocking factor and within-patient correlation was estimated using *duplicateCorrelation.* All models included sampling position (Near vs Far) as a covariate. Species-level effects were first assessed using a global moderated F-test to identify species-associated heterogeneity, followed by exploratory pairwise comparisons between species. Additional analysis grouped samples by pathogen class to determine differences associated with each class. For class-based analyses, differential expression between pathogen classes was tested using a moderated t-test within the repeated-measures framework. For all analyses, *p*-values were adjusted for multiple testing using the Benjamini–Hochberg false discovery rate (FDR), and proteins with FDR < 0.05 were considered statistically significant. All analyses were conducted in R 4.4.2 using *the tidyverse*, *limma*, and *ggplot2* packages. Package versions are recorded via *renv*.

Immunohistochemistry data were collected as the number of marker-positive cells per unit area (mm^2^) using StrataQuest software. Cell counts in the three position ranges (0–100 µm, 101–200 µm, and ≥201 µm) were converted to sample-level proportions to account for differences in total counts. Repeated measures were handled using mixed-effects models. Proportional data were analysed as compositional data using additive log-ratio transformations, with the ≥201 µm range as the reference. A pseudocount of 0.5 was added to facilitate modelling. Linear mixed-effects models using the *lme4* package in R were fitted for each log-ratio outcome, with class as a fixed effect and patient as a random intercept. Class effects were assessed using likelihood-ratio tests, and a global test of compositional differences was obtained by combining log-ratio tests using Fisher’s method. Model-estimated mean proportions were obtained using the *emmeans* package by back-transformation for visualisation and interpretation. Data were graphically represented using the *ggplot2* package.

## Results

### Study population

The overall study design is shown in **Fig. 1A**. Of the 28 patients initially recruited to the study, 11 were selected for this analysis based on the ease of grain identification in H&E sections. Their demographic, clinical, and histological characteristics, and causal organisms are shown in Table 1. None of the included patients had prior medical or surgical treatment. Representative histopathology (H&E) images are provided in **Fig. S1**. Causative organisms were *Actinomadura pelletieri* (P1, 4, 7, and 23), *Streptomyces somaliensis* (P2, 25) and *Madurella mycetomatis* (P8, 17, 24, 27, and 28).

**Figure 1:**
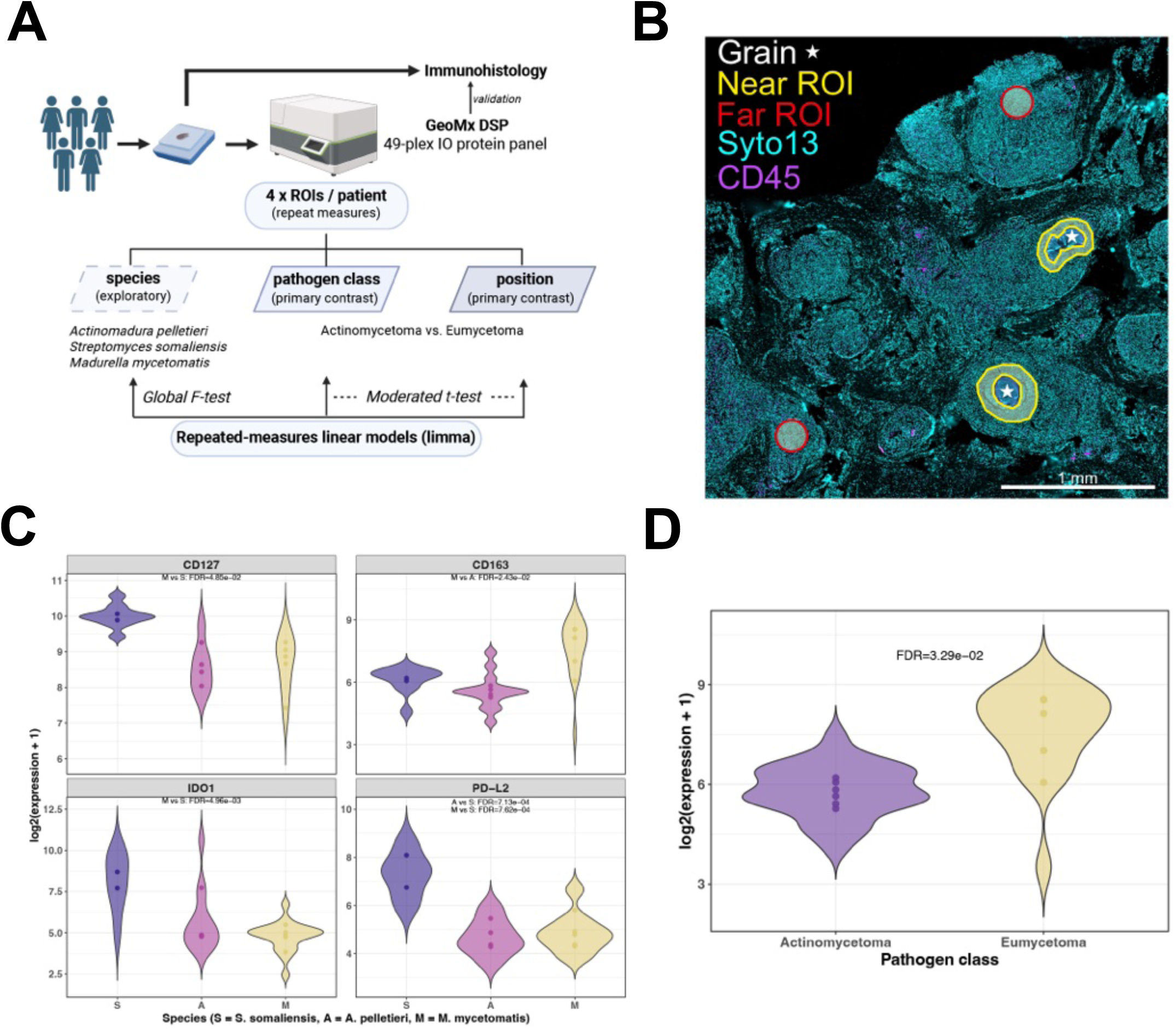
Immune microenvironment of mycetoma lesions. **A.** Experimental workflow for spatial profiling mycetoma lesions using Digital Spatial Profiling GeoMx assay (DSP), with validation by immunohistology. **B.** Representative section showing placement of ROIs for DSP analysis. Grain indicated by a star. **C.** Violin plots showing expression of CD127, CD163, IDO1 and PD-L2 in patients with mycetoma caused by *S. somaliensis*, *A. pelletieri* and *M. mycetomatis*. FDR for comparisons is shown in the Figure. Points represent means value for each patient. **D.** Violin plot showing CD163 expression by mycetoma pathogen class. In C and D, significance was determined using a moderated t-test with correction for multiple testing and repeated measures. Significant FDRs are shown in each panel. Panel A was created in BioRender. Kaye, P. (2026) https://BioRender.com/ga0aq4s.

**Table 1.** Details the clinical characteristics of the eumycetoma and actinomycetoma patients.

| Type | Sex (M/F) | Age (years) | Organism | Lesion size (cm) | Duration (years) | Biopsy site | Histology (grade) |
| --- | --- | --- | --- | --- | --- | --- | --- |
| **Actinomycetoma** |  |  |  |  |  |  |  |
| P1 | M | 16-20 | *A. pelletieri* | >10 | 2 | foot | I + II |
| P2 | F | 26-30 | *S. somaliensis* | 0-10 | 8 | foot | I + II |
| P4 | M | 81-85 | *A. pelletieri* | >10 | 4 | leg | II + III |
| P7 | M | 21-25 | *A. pelletieri* | 0-10 | 2 | foot | II + III |
| P23 | M | 66-70 | *A. pelletieri* | 0-10 | 2 | foot | III |
| P25 | M | 36-40 | *S. somaliensis* | >10 | 1 | foot | III |
| **Eumycetoma** |  |  |  |  |  |  |  |
| P8 | M | 51-55 | *M. mycetomatis* | >10 | 8 | leg | I + II |
| P17 | M | 21-25 | *M. mycetomatis* | 0-10 | 3 | hand | II |
| P24 | M | 16-20 | *M. mycetomatis* | 0-10 | 1 | hand | I + II |
| P27 | M | 26-30 | *M. mycetomatis* | 0-10 | 2 | leg | II + III |
| P28 | M | 41-45 | *M. mycetomatis* | >10 | 10 | leg | II + III |

\*Histopathology: classification based on [16]

### Spatial profiling of mycetoma surgical biopsies

For these 11 patients, we performed spatial protein profiling. For each patient, we examined 4 regions of interest (ROIs) based on their positions relative to grains: two surrounding grains (“Near”) and two in tissue areas devoid of grains (“Far”) (**Fig. 1B**). From an initial protein panel comprising 43 immune protein targets, 32 were expressed, passed QC and thresholding (see Methods) and were used for further analysis (**Fig. S2** and **Table S1**). Protein counts were normalised to nuclei, allowing estimates of per-cell protein expression independent of the extent of cellular infiltration.

As a first exploratory analysis, we assessed per-cell expression of immune proteins based on causative species and independently of position. Protein expression data were analysed using linear models that accounted for repeated measures within each patient and sampling position. For all 32 proteins, there was evidence of an overall species effect (FDR < 0.05), suggesting species-associated immune response heterogeneity (**Fig. S3**). However, when direct pairwise comparisons were performed between individual species, only four proteins showed statistically significant species-related effects (CD127, CD163, IDO1, PD-L2; **Fig.1C**). This result is consistent with limited power due to the small sample size and unequal patient numbers (n = 2, 4, and 5).

To partially mitigate these limitations, and because these species belong to the two major classes of pathogens (shaping potentially different immune responses), we grouped samples into actimomycetoma (*S. somaliensis* and *A. pellettieri*; n = 6) and eumycetoma (*M. mycetomatis*; n = 5) to examine response differences at this broader taxonomic level. Using a repeated-measures model similarly adjusted for position, only CD163 expression was significantly different, with greater per-cell expression in eumycetoma patients compared to actinomycetoma patients (FDR = 0.033; **Fig 1D**). CD163 is a high-affinity scavenger receptor commonly associated with M2 macrophage activation, immunosuppression and resistance to immunotherapy [35].

In contrast to these observed species- and pathogen class-associated effects, sampling position (i.e., Near vs Far) was more robustly associated with differences in protein expression across all modelling strategies (**Fig. 2A**). Near ROIs showed significantly greater expression of leucocyte (CD45) and lineage (CD3, CD8, CD56, CD66b, panCK) markers, as well as leucocyte-associated activation and immunoregulatory proteins (CD45RO, ARG1, CTLA4, IDO1, and VISTA). In contrast to this enrichment in immune-related proteins in near ROIs, far ROIs were enriched for SMA, Fibronection and CD34, indicative of a more stromal-rich environment (**Fig. 2A**). Pairwise correlation analysis for the subset of proteins significantly associated with near ROIs identified distinct peri-grain niches, notably with high per cell expression of i) CD45, CD3 and CD8 (**Fig. 2B**) and ii) CD66b, ARG1 and VISTA (**Fig. 2B-D**). These niches were less evident in the analysis of the same protein set in far ROIs, though strong correlations between CD3 and CD8 and between CD66b and ARG1 were maintained (**Fig. S4A-C**). VISTA remained correlated with CD66b, but was more closely correlated with proteins defining a lymphoid niche (**Fig S4A**). Given that ARG1 and VISTA have been identified in recent years as major contributors to immunosuppression within the tumour microenvironment [36–38], these data indicate that the microenvironment near grains can be characterised as “immune hot”, with evidence of an immunosuppressive microenvironment.

**Figure 2.**
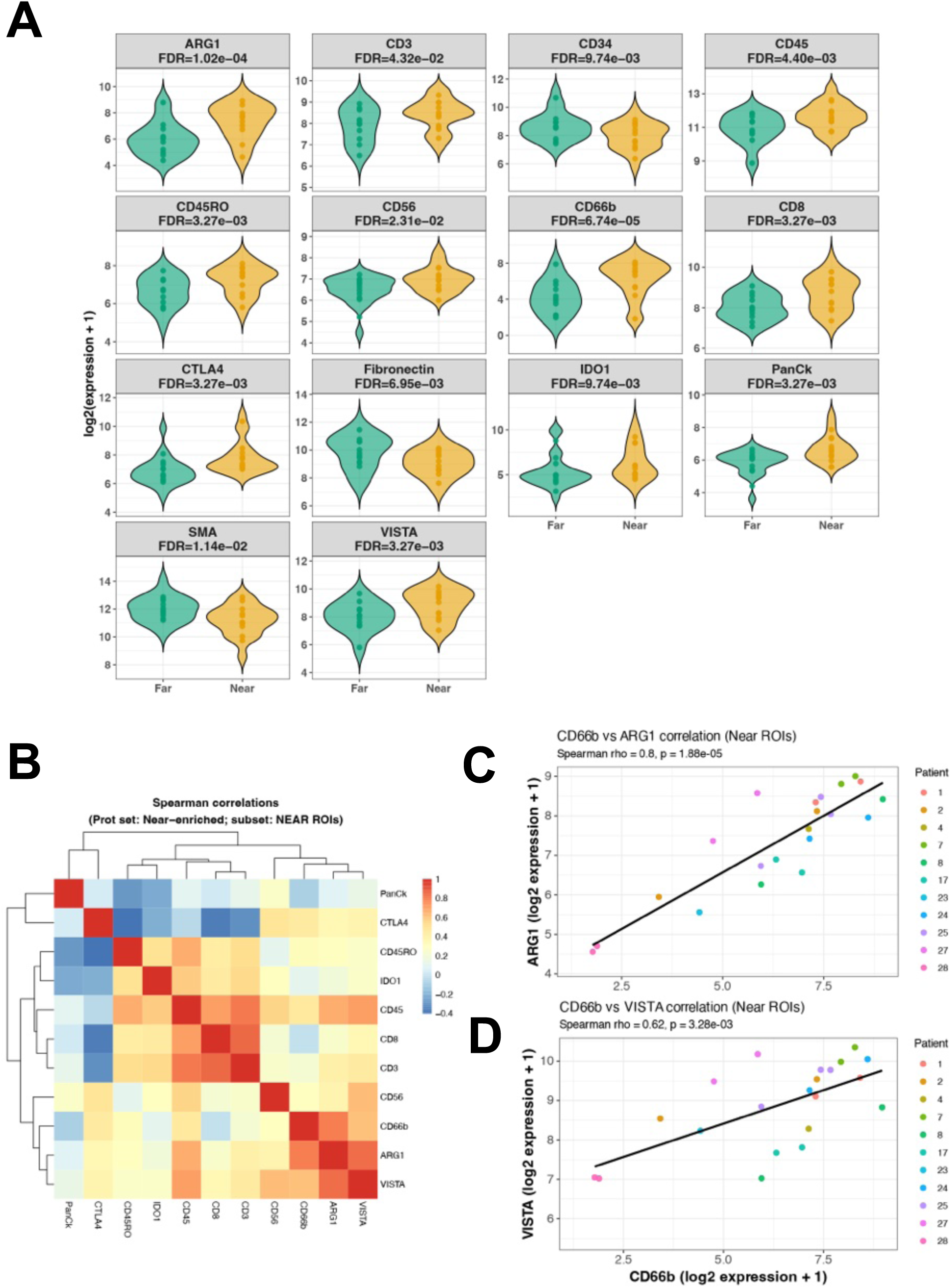
Per cell protein expression in near and far ROIs. **A.** Violin plots showing protein expression for 14 targets showing significant differences in expression between near and far ROIs. Significance was determined using a moderated t-test with correction for multiple testing and repeated measures. **B.** Spearman correlation of 11 near-associated proteins in near ROIs. **C, D.** Correlation between CD66b and ARG1 (C) and VISTA (D) in near ROIs, indicating individual patients by colour.

### CD66b^+^ cells accumulate near grains

CD66b^+^ cells expressing ARG1 and/or VISTA have previously been associated with the generation of an immunosuppressive tumour microenvironment [37, 39]. To confirm that CD66b^+^ cells were preferentially localised near grains, we stained tissue sections with antibodies directed to CD3, CD68 and CD66b. This analysis excluded P21, where material was limited, but included two additional actimomycetoma samples (P12 and P23) where grains were eventually found in FFPE sections. Qualitative assessment indicated that CD66b^+^ as well as CD3^+^ cells were more commonly observed in close proximity to grains compared to other regions of the tissue, whereas CD68^+^ macrophages were more diffusely located (**Fig 3A, top panels**).

**Figure 3.**
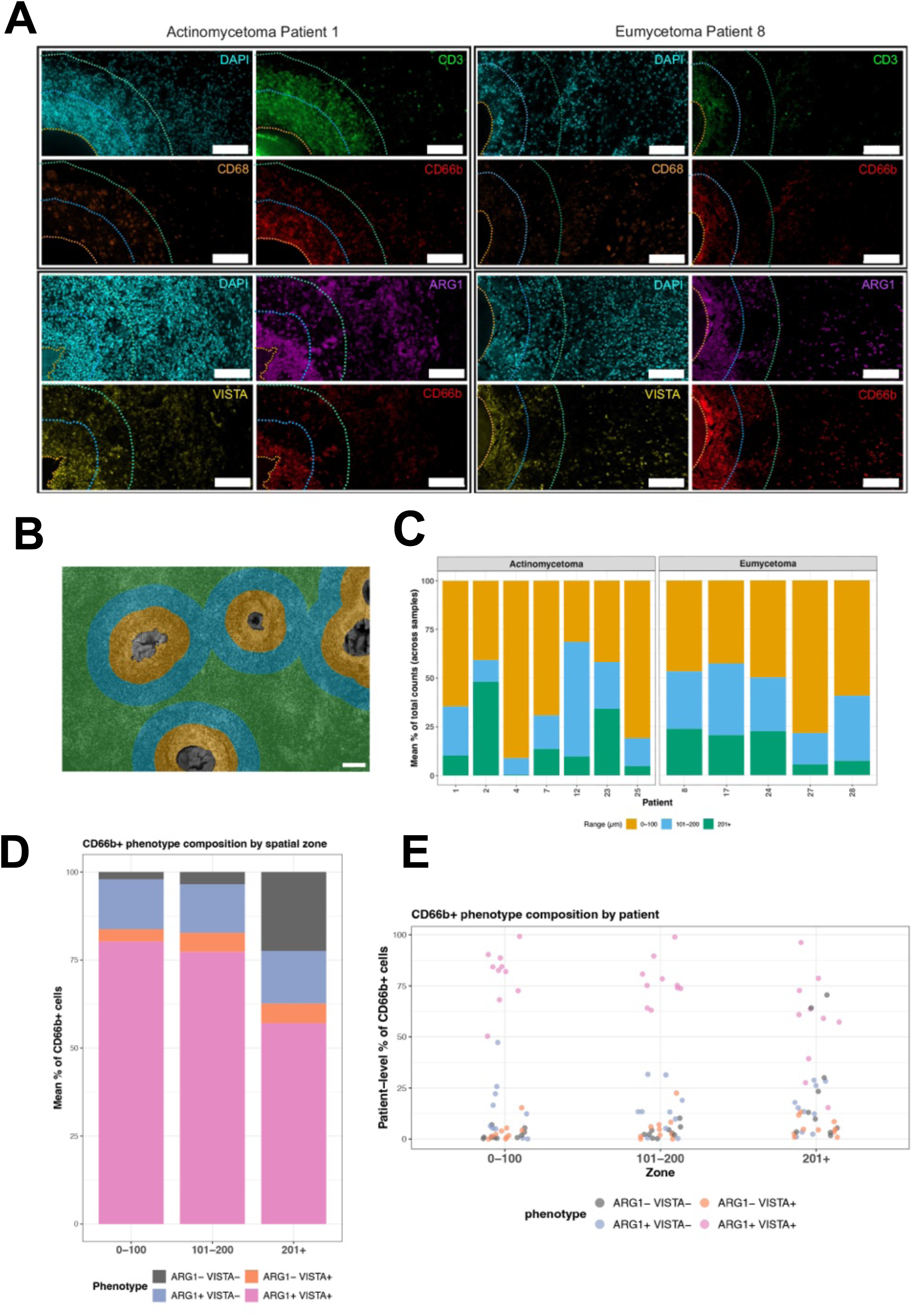
Distribution and expression of ARG1 and VISTA on CD66b^+^ cells. **A.** Representative immunostaining for P1 and P8 showing in top panels DAPI (blue) CD3 (green), CD68 (orange) and CD66b (red) and in bottom panels DAPI (blue) ARG1 (purple), VISTA (yellow) and CD66b (red). The grain boundary is located to the left of the image (orange dotted line) with 0–100 µm and 101–200 µm zone boundaries marked by blue and green dotted lines, respectively. Scale bar = 100 µm. **B.** Representative image to show zone based analysis in Stratquest. Scale bar = 100 µm. **C.** Stacked bar plots show patient-level mean proportions of counts in three zones (0–100 µm, yellow; 101–200 µm, blue; ≥201 µm, green), averaged across repeated samples within each patient. Bars are faceted by class. Mixed-effects compositional analysis (see Methods). **D, E.** Phenotypic composition of CD66b^+^ cells by zone, shown for all patients (D) and individually by patient (E). Legend as for C. Global test, Fisher’s method: χ² = 47.6, p = 1.4 × 10⁻⁸.

To quantify the distribution of CD66b^+^ cells relative to grain perimeters, we created ROIs (zones) extending 0-100 μm, 101-200 μm, and 200+ μm from grain boundaries (**Fig. 3A, B**), To assess the spatial distribution of CD66b⁺ cells across zones, cell densities (cells/mm²) were converted to proportions within each sample and analysed as compositional data using additive log-ratio transformation and mixed-effects modelling with patient as a random effect. This analysis indicated that CD66b^+^ cells were preferentially distributed closest to grains but this was not significantly different between eumycetoma and actinomycetoma patients (p = 0.79, χ^2^ = 1.68; Global Fisher’s test; **Fig 3C**). Back transformed model-estimated compositions indicated that CD66b⁺ cells were preferentially distributed towards the 0–100 µm and 101–200 µm zones relative to the 200+ µm zone, with estimated enrichments of 6.8 and 4.1-fold and 2.3- and 2.1-fold, for actinomycetoma and eumycetoma respectively.

As both ARG1 and VISTA can be expressed by CD66b^+^ cells as well as tumour-associated macrophages ( Pasupuleti et al., 2021) and some T cells [40], we co-stained sections for CD66b, ARG1, and VISTA to confirm whether these molecules were co-expressed (**Fig. 3A, bottom panels**). CD66b^+^ cell phenotype composition was significantly affected by spatial position (p = 1.4 x10^-8^, χ² = 47.6; Global Fisher’s test; **Fig. 3D, E**). In the most proximal zone to grains, almost all CD66b^+^ cells expressed ARG1 and ∼80% coexpressed ARG1 and VISTA. No significant compositional difference was observed between 0–100 µm and 101–200 µm ranges ( p= 0.28) whereas both proximal zones were significantly different from the 200+ µm zone (p < 0.0001 and p < 0.0005, respectively). These changes were driven by relative enrichment of CD66b^+^ARG1^+^VISTA^+^ cells. Collectively, these results indicate that spatial proximity to a grain is associated with a distinct CD66b^+^ cell phenotype characterised by co-expression of ARG1 and VISTA. The concordance between DSP protein expression (**Fig. 2A**), marker correlation (**Fig. 2B-D**), and single-cell immunophenotyping (**Fig. 3**) supports the presence of a spatially organised, immunoregulatory CD66b^+^ARG1^+^VISTA^+^ cell niche that may contribute to local immunosuppression.

## Discussion

Mycetoma is a chronic progressive granulomatous inflammatory disease caused by phylogenetically dissimilar bacterial (actinomycetoma) and fungal (eumycetoma) pathogens. Actinomycetoma is generally considered more aggressive, inflammatory, destructive, and invasive, yet it responds better to a combination of chemotherapeutic agents. Eumycetoma has a more benign behaviour, is less destructive and more localised, but responds poorly to antifungal agents [2, 27, 33]. The data reported here suggest a common pathogenic pathway in mycetoma linked to the recruitment and retention of immunosuppressive CD66b^+^ cells within a peri-grain niche that bears similarities to the tumour microenvironment.

Our analysis at the species and class level indicated minimal differences in expression of immune oncology targets between these two forms of disease. Weakly significant differences in protein expression were observed across all targets, but most did not meet the FDR threshold when comparing species or classes pairwise. The exception was CD163, a marker associated with resident non-migratory M2-like macrophages in the tumour microenvironment [35, 41], which was more highly expressed in eumycetoma patients. Our data emphasise the need for larger cohort studies to ascertain species and class-level differences in immune marker expression. For example, a cohort of 100 patients was used to identify species- and class-specific differences in MMP9 and IL-17A expression [23]. However, such sample sizes are beyond what is practical or generally affordable when employing highly multiplexed spatial technologies.

CD66b is a GPI-anchored glycoprotein constitutively expressed by human neutrophils and other granulocytes, that is upregulated during activation-induced exocytosis of neutrophil granules [42, 43]. CD66b^+^ neutrophils have been identified in the tumour microenvironment where they play an immunoregulatory role [37, 39]. Our finding of increased numbers of leucocytes and, in particular, CD66b^+^ cells near to mycetoma grains is in keeping with previous reports [15, 16, 22], but the first to co-localise expression of the immunosuppressive molecules ARG1 and VISTA to CD66b^+^ cells. Although commonly used as a marker of activated neutrophils, CD66b is also highly expressed on myeloid-derived suppressor cells (MDSCs). As our protein panels do not distinguish MDSCs from neutrophils, we cannot currently say whether ARG1 and VISTA are co-expressed only on neutrophils, on MDSC, or on both. Further studies are therefore required to dissect the functionality and plasticity of mycetoma-associated CD66b^+^ cells and to define the roles of neutrophils and MDSC subpopulations in mycetoma.

Arginase-1 (ARG1) and V-domain Ig Suppressor of T cell Activation (VISTA; PD-1H) have been shown to negatively influence immune effector responses at the tumour site, acting respectively as metabolic and cell membrane-expressed immune checkpoint molecules. In the tumour microenvironment, ARG1 overexpression can inhibit T cell proliferation by depleting arginine [44, 45]. VISTA is a B7-family immune checkpoint molecule constitutively expressed on myeloid cells that ligates T cell-expressed P-selectin glycoprotein ligand-1 (PSGL-1) [46] and leucine-rich repeats and immunoglobulin-like domains 1 (LRIG1) to deliver an inhibitory signal. Both ARG1 and VISTA are expressed in the *Mycobacterium tuberculosis* granuloma [47, 48], and ARG1 expression associated with macrophage polarisation has been observed in the *Schistosoma mansoni* granuloma [49]. Further research into the expression and function of these immunosuppressive molecules in the broader context of granulomatous inflammation is clearly warranted.

The identification of immune checkpoint molecules in the grain microenvironment opens the possibility of targeted adjunct therapy. Novel therapeutics have been identified that target arginase [37, 39, 50–52] and VISTA [36, 53–55]. In cancer immunotherapy, numerous clinical trials have assessed the efficacy of combination chemotherapy with checkpoint inhibitors, aiming to achieve greater therapeutic benefit by reactivating local T cell immunity. Numerous studies have also identified synergies between immune effector mechanisms and antimicrobials, involving both innate (e.g., MAIT cells [56]) and conventional lymphocyte responses [34]. In the case of mycetoma, therefore, there is a compelling argument that alleviating local immunosuppression may also improve the efficacy of antimicrobials and / or limit their duration of use, thereby reducing antimicrobial resistance. This may be particularly true for anti-fungal agents, given the poor prognosis of eumycetoma patients. Hence, whilst our study remains exploratory, our data suggests that a more comprehensive analysis of the immunosuppressive grain-associated microenvironment could lead to the development of novel combination therapies for mycetoma.

Our study has clear limitations. The sample size was small, limiting our analysis of species and class differences to largely exploratory findings. Many of the protein markers in the IO panel were not expressed at sufficiently high levels to be evaluated. Whilst not an uncommon finding [57], this limited the characterisation of the grain-associated microenvironment, notably with regard T cell phenotypes. Improved multiplexed protein profiling approaches and / or single cell spatial transcriptomics may overcome this issue for future studies. Finally, although our ROIs for spatial profiling were relatively large and captured protein expression from multiple cell types, this limitation was mitigated by orthogonal validation at single cell level by quantitative immunohistology.

In conclusion, this study showed similar immune protein expression patterns and cellular infiltrates in the mycetoma-immune microenvironment for both eumycetoma and actinmycetoma patients, with a clear grain-associated microenvironment characterised by the enhanced presence of CD66b^+^ARG1^+^VISTA^+^ myeloid cells. Given the clinical challenge associated with conventional antimicrobial treatment, further exploration of the therapeutic potential of modulators of the grain-associated microenvironment could be pursued.

## Acknowledgements

The authors thank Salma Ramadan and Emanwell Master for technical assistance and the patients and their families for providing samples for this study.

## Data availability statement

Raw data files for DSP and immunohistochemistry are included as supplementary tables in this manuscript. Processed count data and code for statistical analysis and plotting are provided in a GitHub repository at https://github.com/paulkaye/mycetomaDSP.github.io

## Author contributions

**Conceptualization**: AHF, PMK, AMM, MO

**Data curation:** HA, PMK, MO.

**Formal analysis:** HA, PMK, MO

**Funding acquisition:** AHF, PMK, MO

**Investigation:** HA, SMB, GC, POT, MO

**Methodology:** HA, GC, PMK, POT, MO

**Resources:** AHF, AMM, PMK

**Supervision:** PMK

**Writing** – original draft: PMK, MO

**Writing** – review & editing: all authors

## Supplementary Tables and Figures

**Table S1. DSP raw data file**

**Fig. S1. Representative histology of mycetoma patients (H&E)**

**Fig. S2. GeoMx protein expression across all ROIs.**

**Fig. S3. Protein expression by species**

**Fig. S4. Correlations plots for Far ROIs**

## References

1. Fahal AH. Mycetoma: A global medical and socio-economic dilemma. PLoS Negl Trop Dis. 2017;11(4):e0005509. Epub 20170420. doi: 10.1371/journal.pntd.0005509. PubMed PMID: 28426654; PubMed Central PMCID: PMCPMC5398501.

2. Fahal AH, Suliman SH, Hay R. Mycetoma: The Spectrum of Clinical Presentation. Trop Med Infect Dis. 2018;3(3). Epub 20180904. doi: 10.3390/tropicalmed3030097. PubMed PMID: 30274493; PubMed Central PMCID: PMCPMC6161195.

3. Hay RJ, Fahal AH. Mycetoma: an old and still neglected tropical disease. Trans R Soc Trop Med Hyg. 2015;109(3):169–70. doi: 10.1093/trstmh/trv003. PubMed PMID: 25667234.

4. Ahmed AO, van Leeuwen W, Fahal A, van de Sande W, Verbrugh H, van Belkum A. Mycetoma caused by Madurella mycetomatis: a neglected infectious burden. Lancet Infect Dis. 2004;4(9):566–74. doi: 10.1016/S1473-3099(04)01131-4. PubMed PMID: 15336224.

5. Lopez-Martinez R, Mendez-Tovar LJ, Bonifaz A, Arenas R, Mayorga J, Welsh O, et al. [Update on the epidemiology of mycetoma in Mexico. A review of 3933 cases]. Gac Med Mex. 2013;149(5):586–92. PubMed PMID: 24108347.

6. van de Sande WW. Global burden of human mycetoma: a systematic review and meta-analysis. PLoS Negl Trop Dis. 2013;7(11):e2550. Epub 20131107. doi: 10.1371/journal.pntd.0002550. PubMed PMID: 24244780; PubMed Central PMCID: PMCPMC3820768.

7. de Hoog GS, van Diepeningen AD, Mahgoub el S, van de Sande WW. New species of Madurella, causative agents of black-grain mycetoma. J Clin Microbiol. 2012;50(3):988–94. Epub 20120111. doi: 10.1128/JCM.05477-11. PubMed PMID: 22205798; PubMed Central PMCID: PMCPMC3295137.

8. van Belkum A, Fahal A, van de Sande WW. Mycetoma caused by Madurella mycetomatis: a completely neglected medico-social dilemma. Adv Exp Med Biol. 2013;764:179–89. doi: 10.1007/978-1-4614-4726-9_15. PubMed PMID: 23654067.

9. Fahal AH. Mycetoma: a thorn in the flesh. Trans R Soc Trop Med Hyg. 2004;98(1):3–11. doi: 10.1016/s0035-9203(03)00009-9. PubMed PMID: 14702833.

10. Sheehan G, Konings M, Lim W, Fahal A, Kavanagh K, van de Sande WWJ. Proteomic analysis of the processes leading to Madurella mycetomatis grain formation in Galleria mellonella larvae. PLoS Negl Trop Dis. 2020;14(4):e0008190. Epub 20200408. doi: 10.1371/journal.pntd.0008190. PubMed PMID: 32267851; PubMed Central PMCID: PMCPMC7141616.

11. van de Sande WW, Fahal AH, Goodfellow M, Mahgoub el S, Welsh O, Zijlstra EE. Merits and pitfalls of currently used diagnostic tools in mycetoma. PLoS Negl Trop Dis. 2014;8(7):e2918. Epub 20140703. doi: 10.1371/journal.pntd.0002918. PubMed PMID: 24992636; PubMed Central PMCID: PMCPMC4080999.

12. Zijlstra EE, van de Sande WWJ, Welsh O, Mahgoub ES, Goodfellow M, Fahal AH. Mycetoma: a unique neglected tropical disease. Lancet Infect Dis. 2016;16(1):100–12. doi: 10.1016/S1473-3099(15)00359-X. PubMed PMID: 26738840.

13. Ahmed SA, van den Ende BH, Fahal AH, van de Sande WW, de Hoog GS. Rapid identification of black grain eumycetoma causative agents using rolling circle amplification. PLoS Negl Trop Dis. 2014;8(12):e3368. Epub 20141204. doi: 10.1371/journal.pntd.0003368. PubMed PMID: 25474355; PubMed Central PMCID: PMCPMC4256478.

14. HS EL, Mahgoub E, Mahmoud N, Fahal AH. Use of immunoblotting in testing Madurella mycetomatis specific antigen. Trans R Soc Trop Med Hyg. 2016;110(5):312–6. doi: 10.1093/trstmh/trw023. PubMed PMID: 27198216; PubMed Central PMCID: PMCPMC4914873.

15. el Hassan AM, Fahal AH, Ahmed AO, Ismail A, Veress B. The immunopathology of actinomycetoma lesions caused by Streptomyces somaliensis. Trans R Soc Trop Med Hyg. 2001;95(1):89–92. doi: 10.1016/s0035-9203(01)90346-3. PubMed PMID: 11280076.

16. Fahal AH, el Toum EA, el Hassan AM, Mahgoub ES, Gumaa SA. The host tissue reaction to Madurella mycetomatis: new classification. J Med Vet Mycol. 1995;33(1):15–7. PubMed PMID: 7650573.

17. Hoeksema MA, Scicluna BP, Boshuizen MC, van der Velden S, Neele AE, Van den Bossche J, et al. IFN-gamma priming of macrophages represses a part of the inflammatory program and attenuates neutrophil recruitment. J Immunol. 2015;194(8):3909–16. Epub 20150306. doi: 10.4049/jimmunol.1402077. PubMed PMID: 25750432.

18. Ibrahim AI, El Hassan AM, Fahal A, van de Sande WW. A histopathological exploration of the Madurella mycetomatis grain. PLoS One. 2013;8(3):e57774. Epub 20130306. doi: 10.1371/journal.pone.0057774. PubMed PMID: 23483927; PubMed Central PMCID: PMCPMC3590280.

19. Nasr A, Abushouk A, Hamza A, Siddig E, Fahal AH. Th-1, Th-2 Cytokines Profile among Madurella mycetomatis Eumycetoma Patients. PLoS Negl Trop Dis. 2016;10(7):e0004862. Epub 20160719. doi: 10.1371/journal.pntd.0004862. PubMed PMID: 27434108; PubMed Central PMCID: PMCPMC4951069.

20. Abushouk A, Nasr A, Masuadi E, Allam G, Siddig EE, Fahal AH. The Role of Interleukin-1 cytokine family (IL-1beta, IL-37) and interleukin-12 cytokine family (IL-12, IL-35) in eumycetoma infection pathogenesis. PLoS Negl Trop Dis. 2019;13(4):e0007098. Epub 20190404. doi: 10.1371/journal.pntd.0007098. PubMed PMID: 30946748; PubMed Central PMCID: PMCPMC6483278.

21. Fahal A, Mahgoub el S, El Hassan AM, Abdel-Rahman ME, Alshambaty Y, Hashim A, et al. A new model for management of mycetoma in the Sudan. PLoS Negl Trop Dis. 2014;8(10):e3271. Epub 20141030. doi: 10.1371/journal.pntd.0003271. PubMed PMID: 25356640; PubMed Central PMCID: PMCPMC4214669.

22. Guimaraes CC, Castro LG, Sotto MN. Lymphocyte subsets, macrophages and Langerhans cells in actinomycetoma and eumycetoma tissue reaction. Acta Trop. 2003;87(3):377–84. doi: 10.1016/s0001-706x(03)00139-6. PubMed PMID: 12875932.

23. Siddig EE, Mohammed Edris AM, Bakhiet SM, van de Sande WWJ, Fahal AH. Interleukin-17 and matrix metalloprotease-9 expression in the mycetoma granuloma. PLoS Negl Trop Dis. 2019;13(7):e0007351. Epub 20190711. doi: 10.1371/journal.pntd.0007351. PubMed PMID: 31295246; PubMed Central PMCID: PMCPMC6622479.

24. Mhmoud NA, Fahal AH, van de Sande WW. The association between the interleukin-10 cytokine and CC chemokine ligand 5 polymorphisms and mycetoma granuloma formation. Med Mycol. 2013;51(5):527–33. Epub 20121204. doi: 10.3109/13693786.2012.745201. PubMed PMID: 23210681.

25. Ali RS, Newport MJ, Bakhiet SM, Ibrahim ME, Fahal AH. Host genetic susceptibility to mycetoma. PLoS Negl Trop Dis. 2020;14(4):e0008053. Epub 20200430. doi: 10.1371/journal.pntd.0008053. PubMed PMID: 32352976; PubMed Central PMCID: PMCPMC7192380.

26. Wadal A, Elhassan TA, Zein HA, Abdel-Rahman ME, Fahal AH. Predictors of Post-operative Mycetoma Recurrence Using Machine-Learning Algorithms: The Mycetoma Research Center Experience. PLoS Negl Trop Dis. 2016;10(10):e0005007. Epub 20161031. doi: 10.1371/journal.pntd.0005007. PubMed PMID: 27798643; PubMed Central PMCID: PMCPMC5087941.

27. Welsh O, Al-Abdely HM, Salinas-Carmona MC, Fahal AH. Mycetoma medical therapy. PLoS Negl Trop Dis. 2014;8(10):e3218. Epub 20141016. doi: 10.1371/journal.pntd.0003218. PubMed PMID: 25330342; PubMed Central PMCID: PMCPMC4199551.

28. Zein HA, Fahal AH, Mahgoub el S, El Hassan TA, Abdel-Rahman ME. Predictors of cure, amputation and follow-up dropout among patients with mycetoma seen at the Mycetoma Research Centre, University of Khartoum, Sudan. Trans R Soc Trop Med Hyg. 2012;106(11):639–44. Epub 20120731. doi: 10.1016/j.trstmh.2012.07.003. PubMed PMID: 22854685.

29. Kloezen W, Meis JF, Curfs-Breuker I, Fahal AH, van de Sande WW. In vitro antifungal activity of isavuconazole against Madurella mycetomatis. Antimicrob Agents Chemother. 2012;56(11):6054–6. Epub 20120910. doi: 10.1128/AAC.01170-12. PubMed PMID: 22964246; PubMed Central PMCID: PMCPMC3486573.

30. van Belkum A, Fahal AH, van de Sande WW. In vitro susceptibility of Madurella mycetomatis to posaconazole and terbinafine. Antimicrob Agents Chemother. 2011;55(4):1771–3. Epub 20110124. doi: 10.1128/AAC.01045-10. PubMed PMID: 21263050; PubMed Central PMCID: PMCPMC3067195.

31. van de Sande WW, de Kat J, Coppens J, Ahmed AO, Fahal A, Verbrugh H, et al. Melanin biosynthesis in Madurella mycetomatis and its effect on susceptibility to itraconazole and ketoconazole. Microbes Infect. 2007;9(9):1114–23. Epub 20070518. doi: 10.1016/j.micinf.2007.05.015. PubMed PMID: 17644456.

32. van de Sande WW, Fahal AH, Bakker-Woudenberg IA, van Belkum A. Madurella mycetomatis is not susceptible to the echinocandin class of antifungal agents. Antimicrob Agents Chemother. 2010;54(6):2738–40. Epub 20100329. doi: 10.1128/AAC.01546-09. PubMed PMID: 20350944; PubMed Central PMCID: PMCPMC2876377.

33. Suleiman SH, Wadaella el S, Fahal AH. The Surgical Treatment of Mycetoma. PLoS Negl Trop Dis. 2016;10(6):e0004690. Epub 20160623. doi: 10.1371/journal.pntd.0004690. PubMed PMID: 27336736; PubMed Central PMCID: PMCPMC4919026.

34. Margolis E, Rosch JW. Fitness Landscape of the Immune Compromised Favors the Emergence of Antibiotic Resistance. ACS Infect Dis. 2018;4(9):1275–7. Epub 20180802. doi: 10.1021/acsinfecdis.8b00158. PubMed PMID: 30070470; PubMed Central PMCID: PMCPMC6358436.

35. van Elsas MJ, Labrie C, Etzerodt A, Charoentong P, van Stigt Thans JJC, Van Hall T, et al. Invasive margin tissue-resident macrophages of high CD163 expression impede responses to T cell-based immunotherapy. J Immunother Cancer. 2023;11(3). doi: 10.1136/jitc-2022-006433. PubMed PMID: 36914207; PubMed Central PMCID: PMCPMC10016286.

36. Gao Y, He Y, Tang Y, Chen ZS, Qu M. VISTA: A Novel Checkpoint for Cancer Immunotherapy. Drug Discov Today. 2024;29(7):104045. Epub 20240524. doi: 10.1016/j.drudis.2024.104045. PubMed PMID: 38797321.

37. Tyrinova TV, Batorov EV, Aristova TA, Ushakova GY, Sizikova SA, Denisova VV, et al. Expression of Inhibitory Molecules (Arginase-1, IDO, and PD-L1) by Myeloid-Derived Suppressor Cells in Multiple Myeloma Patients in Remission. Bull Exp Biol Med. 2022;174(1):71–5. Epub 20221128. doi: 10.1007/s10517-022-05651-8. PubMed PMID: 36437327.

38. Sandstrom Gerdtsson A, Knulst M, Botling J, Mezheyeuski A, Micke P, Ek S. Phenotypic characterization of spatial immune infiltration niches in non-small cell lung cancer. Oncoimmunology. 2023;12(1):2206725. Epub 20230427. doi: 10.1080/2162402X.2023.2206725. PubMed PMID: 37139184; PubMed Central PMCID: PMCPMC10150622.

39. Miret JJ, Kirschmeier P, Koyama S, Zhu M, Li YY, Naito Y, et al. Suppression of Myeloid Cell Arginase Activity leads to Therapeutic Response in a NSCLC Mouse Model by Activating Anti-Tumor Immunity. J Immunother Cancer. 2019;7(1):32. Epub 20190206. doi: 10.1186/s40425-019-0504-5. PubMed PMID: 30728077; PubMed Central PMCID: PMCPMC6366094.

40. West EE, Merle NS, Kaminski MM, Palacios G, Kumar D, Wang L, et al. Loss of CD4(+) T cell-intrinsic arginase 1 accelerates Th1 response kinetics and reduces lung pathology during influenza infection. Immunity. 2023;56(9):2036–53 e12. Epub 20230811. doi: 10.1016/j.immuni.2023.07.014. PubMed PMID: 37572656; PubMed Central PMCID: PMCPMC10576612.

41. Jayasingam SD, Citartan M, Thang TH, Mat Zin AA, Ang KC, Ch’ng ES. Evaluating the Polarization of Tumor-Associated Macrophages Into M1 and M2 Phenotypes in Human Cancer Tissue: Technicalities and Challenges in Routine Clinical Practice. Front Oncol. 2019;9:1512. Epub 20200124. doi: 10.3389/fonc.2019.01512. PubMed PMID: 32039007; PubMed Central PMCID: PMCPMC6992653.

42. Lakschevitz FS, Hassanpour S, Rubin A, Fine N, Sun C, Glogauer M. Identification of neutrophil surface marker changes in health and inflammation using high-throughput screening flow cytometry. Experimental Cell Research. 2016;342(2):200–9. doi: 10.1016/j.yexcr.2016.03.007.

43. Schmidt T, Zündorf J, Grüger T, Brandenburg K, Reiners A-L, Zinserling J, et al. CD66b overexpression and homotypic aggregation of human peripheral blood neutrophils after activation by a gram-positive stimulus. Journal of Leukocyte Biology. 2012;91(5):791–802. doi: 10.1189/jlb.0911483.

44. Czystowska-Kuzmicz M, Sosnowska A, Nowis D, Ramji K, Szajnik M, Chlebowska-Tuz J, et al. Small extracellular vesicles containing arginase-1 suppress T-cell responses and promote tumor growth in ovarian carcinoma. Nature communications. 2019;10(1):3000.

45. Maisonneuve C, Tsang DK, Foerster EG, Robert LM, Mukherjee T, Prescott D, et al. Nod1 promotes colorectal carcinogenesis by regulating the immunosuppressive functions of tumor-infiltrating myeloid cells. Cell Reports. 2021;34(4).

46. Johnston RJ, Su LJ, Pinckney J, Critton D, Boyer E, Krishnakumar A, et al. VISTA is an acidic pH-selective ligand for PSGL-1. Nature. 2019;574(7779):565–70. doi: 10.1038/s41586-019-1674-5.

47. Ashenafi S, Muvva JR, Mily A, Snall J, Zewdie M, Chanyalew M, et al. Immunosuppressive Features of the Microenvironment in Lymph Nodes Granulomas from Tuberculosis and HIV-Co-Infected Patients. Am J Pathol. 2022;192(4):653–70. Epub 20220131. doi: 10.1016/j.ajpath.2021.12.013. PubMed PMID: 35092727; PubMed Central PMCID: PMCPMC9302207.

48. Peyster EG, Smith D, Bittermann T, Bravo PE, Margulies KB. Cardiac sarcoidosis: new insights beyond the granuloma using spatial proteomics. European Heart Journal. 2025:ehaf990. doi: 10.1093/eurheartj/ehaf990.

49. Ye Z, Huang S, Zhang Y, Mei X, Zheng H, Li M, et al. Galectins, Eosinophiles, and Macrophages May Contribute to Schistosoma japonicum Egg-Induced Immunopathology in a Mouse Model. Front Immunol. 2020;11:146. Epub 20200313. doi: 10.3389/fimmu.2020.00146. PubMed PMID: 32231658; PubMed Central PMCID: PMCPMC7082360.

50. Steggerda SM, Bennett MK, Chen J, Emberley E, Huang T, Janes JR, et al. Inhibition of arginase by CB-1158 blocks myeloid cell-mediated immune suppression in the tumor microenvironment. J Immunother Cancer. 2017;5(1):101. Epub 20171219. doi: 10.1186/s40425-017-0308-4. PubMed PMID: 29254508; PubMed Central PMCID: PMCPMC5735564.

51. Naing A, Papadopoulos KP, Pishvaian MJ, Rahma O, Hanna GJ, Garralda E, et al. First-in-human phase 1 study of the arginase inhibitor INCB001158 alone or combined with pembrolizumab in patients with advanced or metastatic solid tumours. BMJ Oncol. 2024;3(1):e000249. Epub 20240509. doi: 10.1136/bmjonc-2023-000249. PubMed PMID: 39886141; PubMed Central PMCID: PMCPMC11235002.

52. Niu F, Yu Y, Li Z, Ren Y, Li Z, Ye Q, et al. Arginase: An emerging and promising therapeutic target for cancer treatment. Biomedicine & Pharmacotherapy. 2022;149:112840. doi: 10.1016/j.biopha.2022.112840.

53. Marin-Acevedo JA, Dholaria B, Soyano AE, Knutson KL, Chumsri S, Lou Y. Next generation of immune checkpoint therapy in cancer: new developments and challenges. J Hematol Oncol. 2018;11(1):39. Epub 20180315. doi: 10.1186/s13045-018-0582-8. PubMed PMID: 29544515; PubMed Central PMCID: PMCPMC5856308.

54. Thisted T, Smith FD, Mukherjee A, Kleschenko Y, Feng F, Jiang ZG, et al. VISTA checkpoint inhibition by pH-selective antibody SNS-101 with optimized safety and pharmacokinetic profiles enhances PD-1 response. Nat Commun. 2024;15(1):2917. Epub 20240404. doi: 10.1038/s41467-024-47256-x. PubMed PMID: 38575562; PubMed Central PMCID: PMCPMC10995192.

55. Martin AS, Molloy M, Ugolkov A, von Roemeling RW, Noelle RJ, Lewis LD, et al. VISTA expression and patient selection for immune-based anticancer therapy. Frontiers in Immunology. 2023;Volume 14 - 2023. doi: 10.3389/fimmu.2023.1086102.

56. Leeansyah E, Boulouis C, Kwa ALH, Sandberg JK. Emerging Role for MAIT Cells in Control of Antimicrobial Resistance. Trends Microbiol. 2021;29(6):504–16. Epub 20201219. doi: 10.1016/j.tim.2020.11.008. PubMed PMID: 33353796.

57. Ashwin H, Milross L, Wilson J, Majo J, Hang Lee JT, Calder G, et al. Identification of a protein expression signature distinguishing early from organising diffuse alveolar damage in COVID-19 patients. J Clin Pathol. 2023;76(8):561–5. Epub 20230309. doi: 10.1136/jcp-2023-208771. PubMed PMID: 36894313; PubMed Central PMCID: PMCPMC10423525.

