## Supplementary figures and images for "A conserved grain-associated immunosuppressive niche in Sudanese patients with mycetoma"

### Supplementary Figure 1

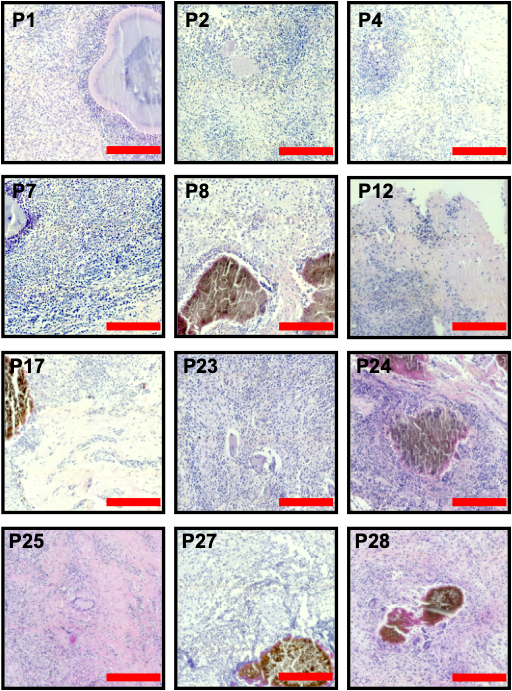

**Figure S1. Representative patient histology (H&E).**

For patient details, see Table S1. Scale bar: 200μm

### Supplementary Figure 3

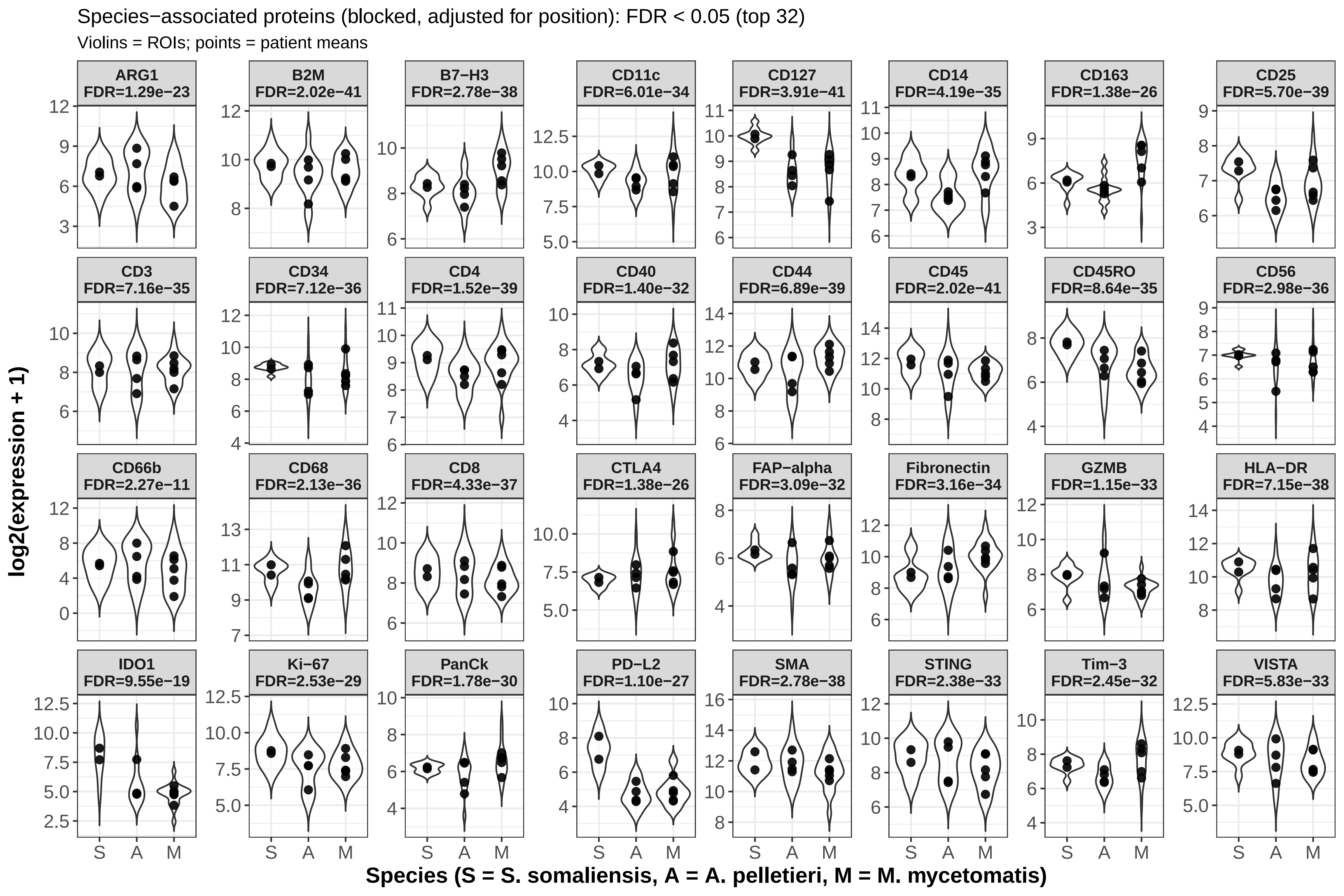
