## Supplementary Figure 2 for "A conserved grain-associated immunosuppressive niche in Sudanese patients with mycetoma"

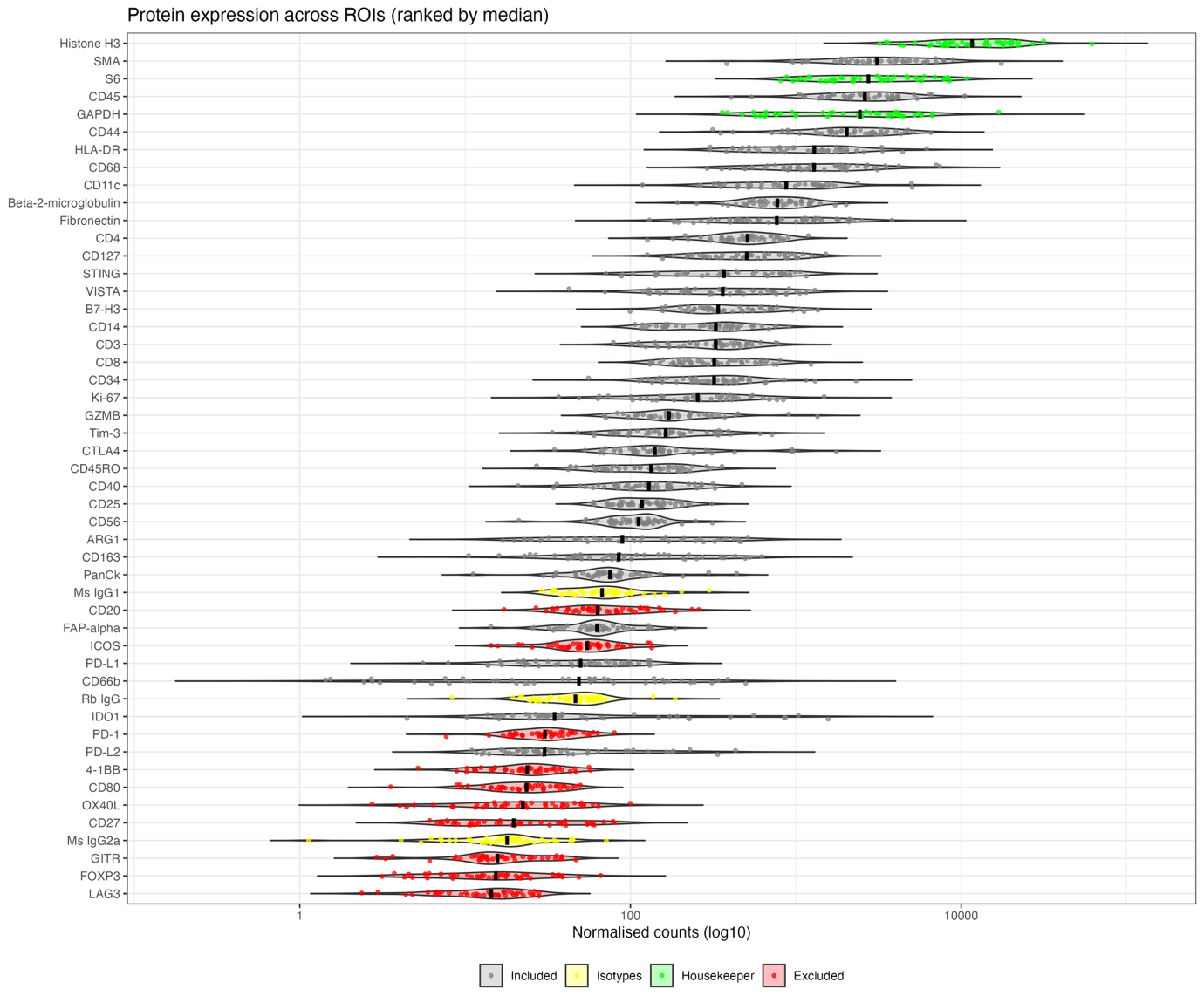

**Figure S2. GeoMx protein expression across all ROIs.**

Data are shown as normalised Log10 counts for 43 immune oncology targets, 3 isotype controls and 3 housekeeping genes. Grey indicates 32 proteins that were included in subsequent analysis as passing QC and thresholding at 3x geomean of isotype controls. Excluded IO proteins (red), isotype controls (yellow) and housekeeping proteins (green) are also indicated. For raw data, see Table S1
