## Supplementary Figure 4 for "A conserved grain-associated immunosuppressive niche in Sudanese patients with mycetoma"

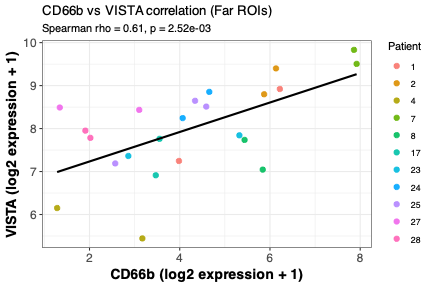

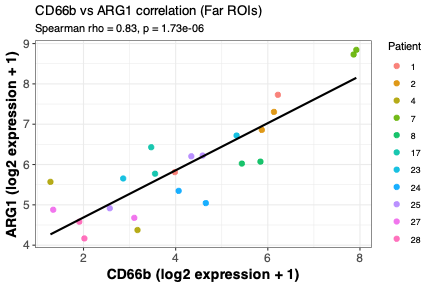

C
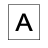

A

B
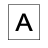

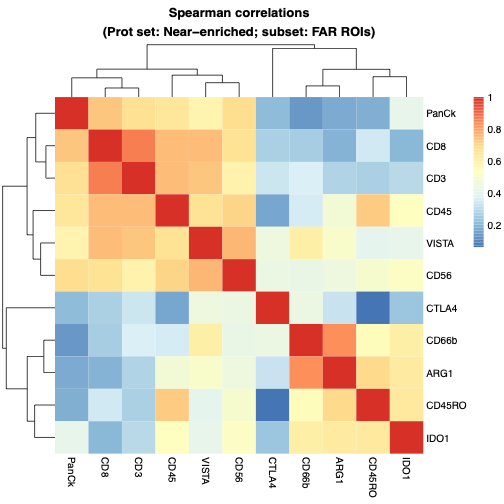

**Figure S4. Correlations plots for Far ROIs**

**A.** Heat map of protein correlation across all Far ROIs using data set enriched in near ROIs. **B, C**. Correlations between CD66b and ARG1 (B) and VISTA (C) in far ROIs.
